# The frequency of mutations in the *pen*A, *mtr*R, *gyr*A and *par*C genes of *Neisseria gonorrhoeae*, the presence of *tet*M gene and antibiotic resistance/susceptibility: a systematic review and meta-analyses

**DOI:** 10.1101/2023.04.26.23289101

**Authors:** Ana Clara Mendes, Renan Pedra de Souza, Diana Bahia

## Abstract

Sexually transmitted infections (STIs) are a major global public health problem and among the most common infectious diseases. Gonorrhoea is an STI that affects only humans, caused by the bacterium *Neisseria gonorrhoeae*. In 2016, the WHO estimated that there were 86.9 million cases of gonorrhoea worldwide. Gonorrhoea is currently one of the most important STIs due to the increased spread and emergence of multiresistant strains. The main goal of this study was to analyse the relationship between resistance or decreased susceptibility to antibiotics in *N. gonorrhoeae* and the presence of mutations in the genes *pen*A, *mtr*R, *tet*M, *gyr*A and *par*C. Therefore, we conducted a systematic review following the PRISMA guidelines. We selected 19 studies for the *pen*A gene, 23 for *gyr*A and *par*C, 18 for *mtr*R, and 12 for *tet*M using the Science Direct and PubMed databases. The included studies addressed susceptibility and resistance to antibiotics and mutations in the selected genes. Then, 21 different meta-analyses of the studies were performed. A meta-analysis was conducted when two or more studies were found for the same gene and the same antibiotic. Meta-analyses of isolates resistant to penicillin, cefixime, and ceftriaxone showed that more than 50% of the isolates had mutations in the genes *pen*A and *mtr*R. Meanwhile, more than 50% of azithromycin-resistant isolates had mutations in the *mtr*R gene, while more than 50% of the isolates resistant and intermediate-resistant to ciprofloxacin had mutations in *gyr*A. Moreover, less than 50% of isolates with intermediate resistance to ciprofloxacin had mutations in *par*C. The plasmid containing the *tet*M gene was found in more than 50% of tetracycline-resistant isolates. Infection monitoring and genetic studies are important to control the spread, which can improve the quality of life of infected individuals and lessen the financial burden on public healthcare systems.

## 1. INTRODUCTION

Sexually transmitted infections (STIs) are one of the main global public health problems and among the main transmissible diseases, affecting the health of men, women, and infants around the world (1). It is estimated that, worldwide, over 1 million new STI cases occur daily (376.4 million new infections annually) due to the four currently treatable STIs: chlamydia, gonorrhoea, syphilis, and trichomoniasis (2). On average, approximately 1 in 25 people has at least one STI. Since the last published data, there has been no significant decline in existing or new infections (1).

Gonorrhoea is an STI that exclusively affects humans and is caused by the bacterium *Neisseria gonorrhoeae*, a Gram-negative diplococcus, also known as gonococcus (3). The infection causes urethritis in males and cervicitis in females. STIs, if unidentified or untreated, can ascend in the genital tract and result in complications (4). In addition, the infection increases the rates of transmission and acquisition of HIV (5). In 2016, the WHO estimated that there were 86.9 million cases of gonorrhoea worldwide. Sexual orientation, sexual behaviour, socioeconomic status, geographical location, culture, and access to sex education are directly related to the epidemiological variety of the disease (1,2).

Gonorrhoea is currently one of the most important STIs due to the increase in the spread and emergence of multidrug-resistant strains; it is also the second most common STI globally (6). For this reason, the WHO has included the bacterium *N. gonorrhoeae* in the list of priorities for combatting transmission and researching new drugs (7). Currently, *N. gonorrhoeae* is resistant to penicillin, tetracycline, macrolides, azithromycin, sulphonamides, quinolones and cephalosporins such as cefixime and ceftriaxone. Many cephalosporin-resistant strains are also resistant to other antibiotics, making *N. gonorrhoeae* a multidrug-resistant bacterium (7,8).

*N. gonorrhoeae* can change its genetic material through several types of mutations, which it uses to adapt and survive in the human host. This species has evolved and acquired or developed mechanisms of resistance to almost all types of antibiotics recommended for use in treatment (8). In addition, *N. gonorrhoeae* is naturally competent for transformation, and transformation occurs more often between *N. gonorrhoeae* and other *Neisseria* species. It has also been shown that commensal *Neisseria* is a reservoir of genetic material for pathogenic *Neisseria* (3,6,9).

When reviewing the antimicrobial resistance mechanisms of *N. gonorrhoea*, Unemo and Shafer (2014) suggested that, due to multidrug resistance, the world may be entering an era of intractable *N. gonorrhoeae*. Antibiotic-resistant *N. gonorrhoeae* can appear as a silent epidemic and the disease and its complications can cause morbidity and economic consequences, as treatment can become more expensive if complications occur. The main genes related to antibiotic resistance of *N. gonorrhoeae* are *pen*A, *bla*TEM, *mtr*R, *tet*M, *gyr*A, and *par*C (10).

Several studies, such as those by Lee et al. (2015) (11) and Calado et al. (2019) (12), assessed minimum inhibitory concentrations (MICs), searched for *N. gonorrhoea* resistance genes, analysed mutations, and related them to resistance or reduced susceptibility to antibiotics. Building on these previous studies, this work involved a systematic review aimed at identifying critical articles focusing on these characteristics and selecting appropriate ones for analysis using a certain set of criteria. The meta-analysis performed here should deepen our understanding of gonorrhoea and help develop solutions to this urgent public health problem.

## 2. METHODS

This systematic review was conducted considering the protocol proposed by PRISMA (MOHER et al., 2009). The phases for the selection of studies were identification, selection, eligibility and inclusion (Table 1). The search focused on the databases PubMed and Science Direct, including all articles within them published to date. The first screening was conducted based on reading of the title and abstract, after which the eligibility was determined through complete reading of the article, with this work being conducted by two researchers independently. This review includes studies on the *Neisseria gonorrhoeae* genes *pen*A, *mtr*R, *tet*M, *par*C and *gyr*A, which are related to resistance, intermediate resistance or reduced susceptibility to penicillin, cefixime, ceftriaxone, azithromycin, ciprofloxacin and tetracycline.

**Table 1.** Phases of the studies selection. The gene *pen*A (*Pen*A *family class A beta-lactamase*) is related to resistance to penicillins and cephalosporins. The genes *gyr*A (*DNA gyrase subunit A)* and *par*C (*DNA topoisomerase IV subunit A)* are related to resistance to quinolones, the *mtr*R (*multidrug efflux system transcriptional repressor)* gene is related to penicillin, tetracycline, cephalosporins and macrolides, and the gene *tet*M (*tetracycline resistance ribosomal protection protein) is related to resistance to tetracyclines*.

| Gene | Identification | Screening |  |  |  |  |
| --- | --- | --- | --- | --- | --- | --- |
|  | Identification from databases | Screened | Excluded | Assessed for eligibility | Excluded | Studies included in qualitative and quantitative analysis |
| *penA* | 226 | 62 | 164 | 19 | 43 | 19 |
| *gyrA* and *parC* | 185 | 53 | 132 | 23 | 30 | 23 |
| *mtrR* | 180 | 66 | 114 | 18 | 48 | 18 |
| *tetM* | 102 | 26 | 76 | 12 | 14 | 12 |

### 2.1 Search Argument

For the PubMed search, the search argument (“Gonorrhea”[Mesh] OR “Gonorrhea”[Title/Abstract] OR “Neisseria gonorrhoeae”[Mesh] OR “Neisseria gonorrhoeae”[Title/Abstract] OR “N. gonorrhoeae”[Title/Abstract])AND (“Antimicrobial resistance”[Title/Abstract] OR “Antimicrobial susceptibility”[Title/Abstract] OR “Resistance Profile”[Title/Abstract] OR “Drug Resistance, Microbial”[Mesh]) AND (“penA”[Title/Abstract]). The argument was adapted for each gene.

For the search in the Science Direct database, the keywords “Neisseria gonorrhoeae, Antimicrobial resistance, *pen*A, *mtr*R, *tet*M, *gyr*A, and *par*C” were used. Filter: research articles.

The inclusion criteria were articles found without the restriction of year and site of publication, both in Portuguese and English and that address susceptibility and resistance to antibiotics and mutations in the chosen genes. The exclusion criteria were articles in languages other than English and Portuguese, articles about other *Nesseria* species, articles that did not analyze antibiotic susceptibility profiles, case report studies, review articles, studies that analyzed the application of a technique, articles that detected susceptibility and mutations in non-clinical isolates, and articles with incomplete genotypic data.

### 2.2 Meta-analysis

The meta-analysis was conducted when two or more studies were found for the same gene and the same antibiotic. The metaprop function of the meta-package was used in the R program (version 4.0.2). The original proportions were combined using the inverse variance method using fixed and random effect models. The significance level was set at 5%.

## 3. RESULTS

### 3.1 Selection of studies

The articles selected for this study are summarized in Table 1.

### 3.2 Studies characteristics

The studies characteristics ware summarized in tables (Table 1 to 6, Supplementary Material) according to the phenotypes. In general, we observed many common characteristics across studies. Included studies investigating antibiotic resistance, decreased susceptibility and mutations in resistance genes started in 1998. Research was conducted in several countries; however, a greater number of studies can be observed in Asian countries, mainly China and Japan (Table 1 to 6, column “Local”). Most of the studies collected samples from urethra, rectum, cervix, endocervix and pharynx and only one study collected samples from other sites such as eye, blood, surgical wound, gastric juice, synovial fluid, and Bartholin abscess. Moreover, the results show a lack of studies in Latin America and Africa. Many studies do not provide details on clinical isolates beyond the study period (second column). Although fewer in number, studies detail gender, sexuality, and sexual behavior. In studies providing sex information, most of the isolates are obtained from samples collected from men, and a minority from samples collected from women.

The studies also reveal which mutations were most found (Table 7 to 10, Supplementary Material). The most common mutations in the *pen*A gene were A501V and mosaic-type mutations. In the *mtr*R gene, deletion of A (adenine), G45D, H105Y, and A39T were the most common mutations. The *gyr*A gene presented the S91F and D95G mutations and the *par*C gene the D86N mutation.

### 3.2 Meta-analysis of studies evaluating penicillin resistance

The results of the meta-analysis for the antibiotic penicillin with the proportion of mutations in the *pen*A and *mtr*R genes are shown in Figure 1 (Supplementary Material). The results of the random model indicate that 87% (CI 42% - 98%) of the analyzed isolates had mutations in the *pen*A gene (Figure 1A). Fixed model results indicate that 95% (CI 86% - 98%) had mutations in the *mtr*R gene (Figure 1B).

**Figure. 1.**
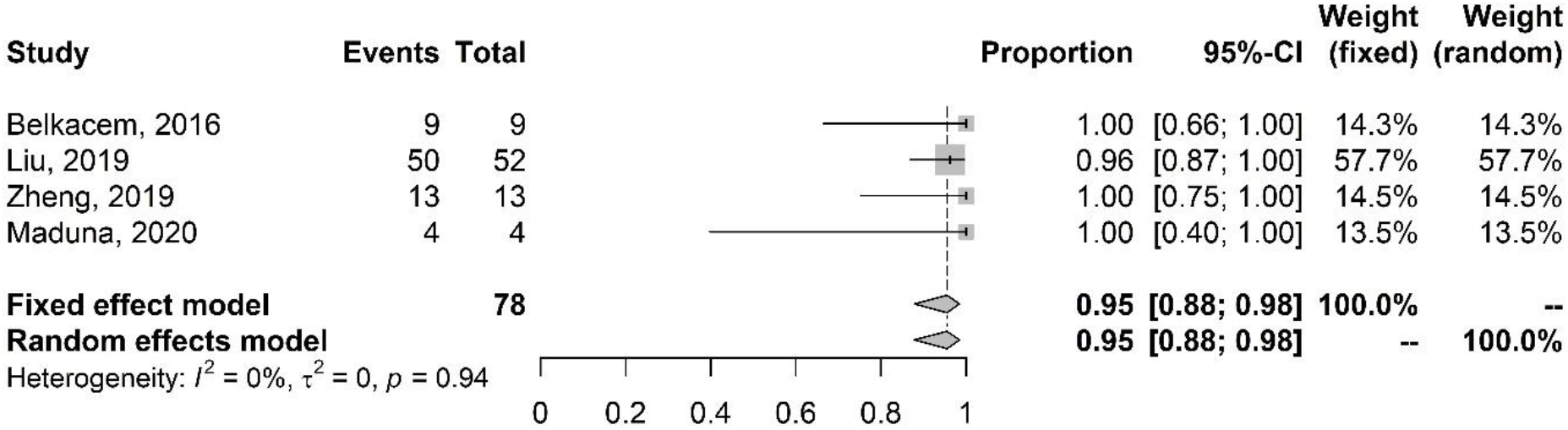
Meta-analysis of the proportion of gene mutations in azithromycin-resistant isolates (*mtr*R gene). The results are presented in forest plots. In its first column (“study”) are the citations of the selected studies. The second column (“events”) contains the number of isolates with mutations in the evaluated gene, while the third column (“total”) shows the total number of samples analyzed in that study. The fourth column presents the proportion estimate followed by their confidence intervals. The sixth and seventh columns present, respectively, the weights for the fixed and random models of the meta-analysis.

The meta-analysis of studies on the antibiotic penicillin with the proportion of mutations in the *mtr*R gene are shown in Figure 2 (Supplementary Material). The results of the fixed model indicate that 85% (CI 71% - 93%) of the analyzed isolates had mutations in the *mtr*R gene.

**Figure. 2.**
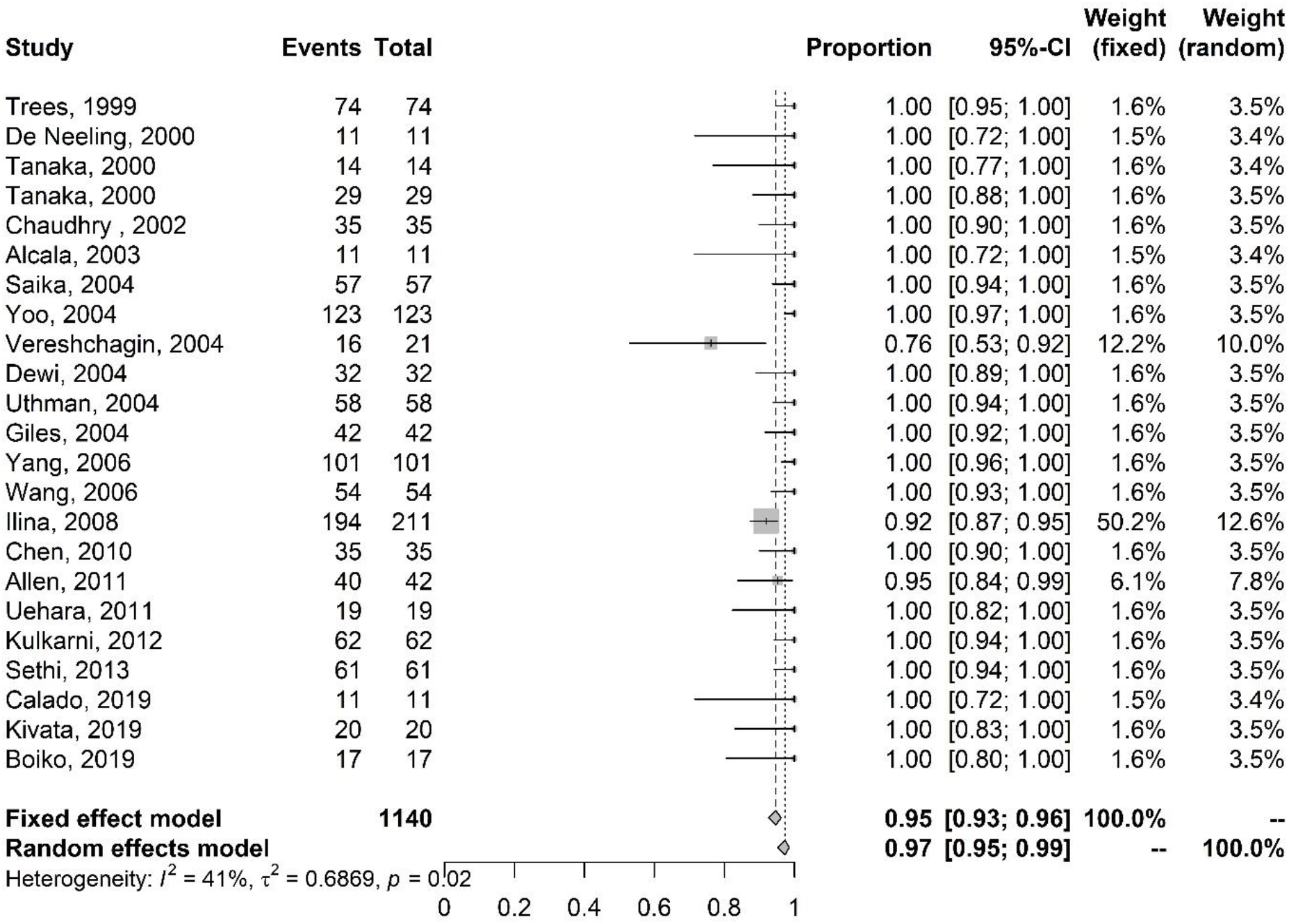
Meta-analysis of the proportion of gene mutations in isolates resistant to ciprofloxacin (*gyr*A gene). The results are presented in forest plots. In its first column (“study”) are the citations of the selected studies. The second column (“events”) contains the number of isolates with mutations in the evaluated gene, while the third column (“total”) shows the total number of samples analyzed in that study. The fourth column presents the proportion estimate followed by their confidence intervals. The sixth and seventh columns present, respectively, the weights for the fixed and random models of the meta-analysis.

### 3.3 Meta-analysis of studies evaluating resistance and reduced susceptibility to cefixime

Figure 3 (Supplementary Material) presents the meta-analysis of studies on cefixime resistance and the proportion of mutations in the *mtr*R and *pen*A genes. The results of the fixed model indicate that 94% (CI 65% - 99%) of the analyzed isolates had mutations in the *mtr*R gene (Figure 3A) and 93% (CI - 77% - 98%) of the isolates had mutations in the *pen*A gene (Figure 3B).

**Figure. 3.**
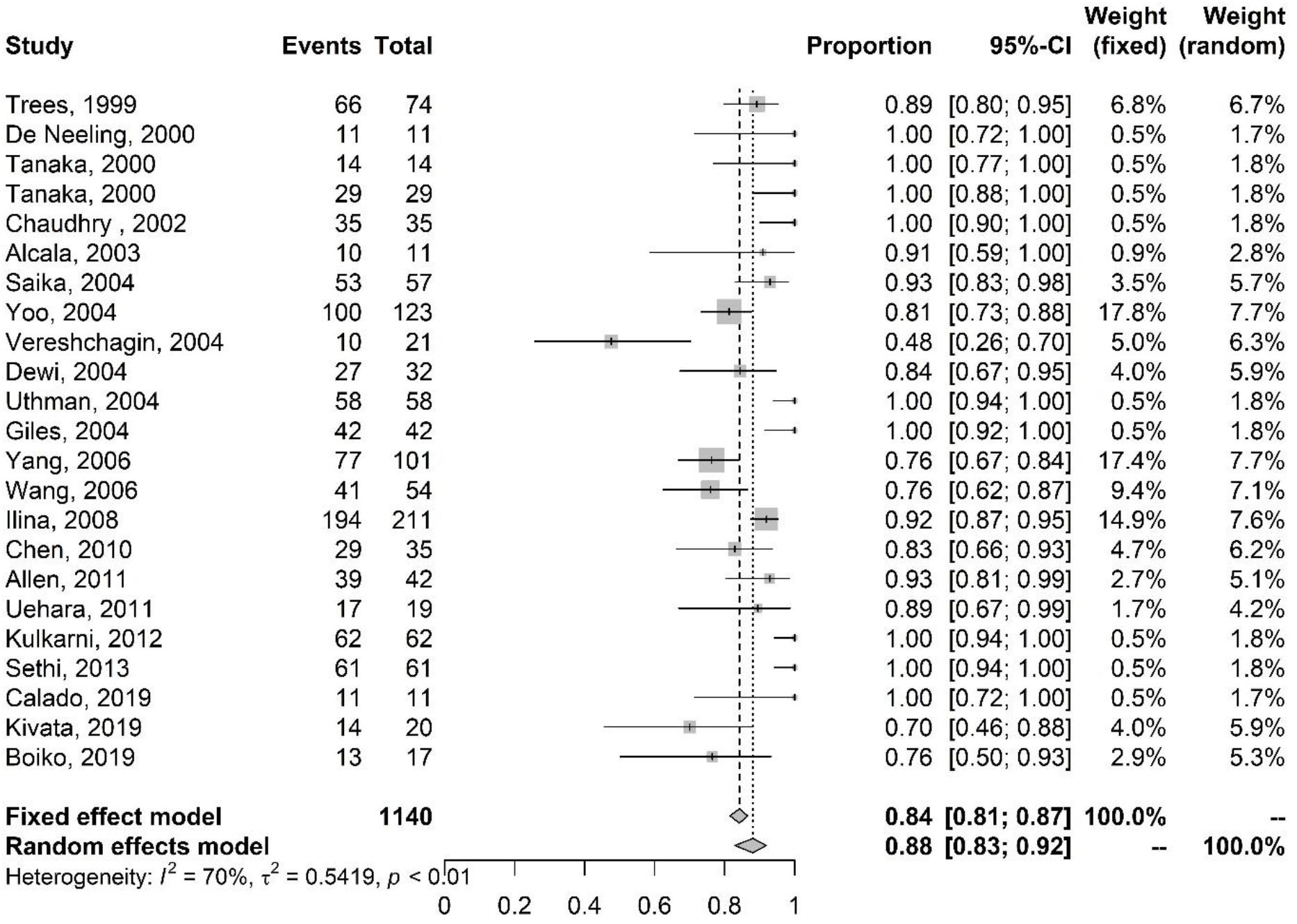
Meta-analysis of the proportion of gene mutations in isolates resistant to ciprofloxacin (parC gene). The results are presented in forest plots. In its first column (“study”) are the citations of the selected studies. The second column (“events”) contains the number of isolates with mutations in the evaluated gene, while the third column (“total”) shows the total number of samples analyzed in that study. The fourth column presents the proportion estimate followed by their confidence intervals. The sixth and seventh columns present, respectively, the weights for the fixed and random models of the meta-analysis.

Figure 4 (Supplementary Material) presents the meta-analysis of studies on reduced susceptibility to cefixime. The results of the random model indicate that 93% (CI 73% - 99%) of the analyzed isolates had mutations in the gene *pen*A (Figure 4A) The results of the fixed model indicate that 96% (CI 88% - 99%) of the isolates had mutations in the *mtr*R gene (Figure 4B).

**Figure. 4.**
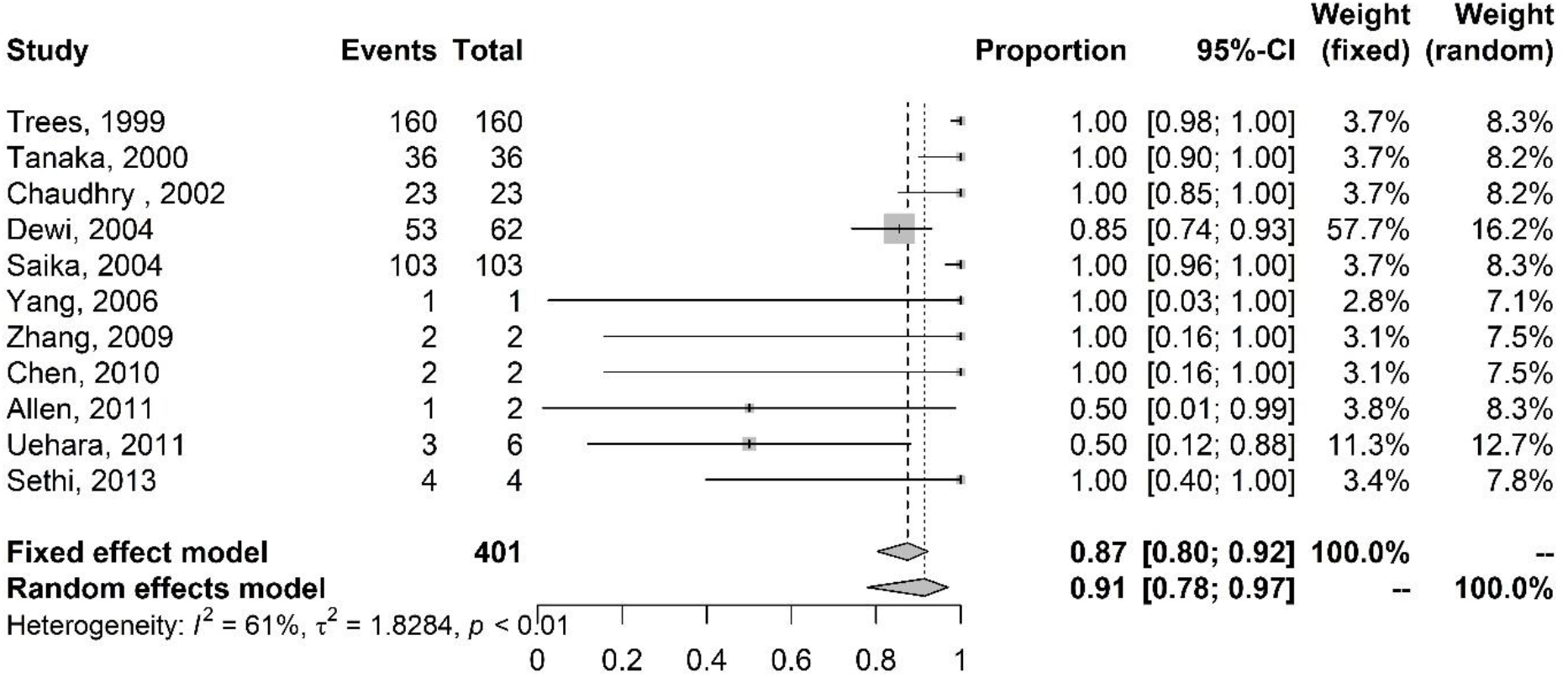
Meta-analysis of the proportion of gene mutations in isolates with intermediate resistance to ciprofloxacin (gyrA gene). The results are presented in forest plots. In its first column (“study”) are the citations of the selected studies. The second column (“events”) contains the number of isolates with mutations in the evaluated gene, while the third column (“total”) shows the total number of samples analyzed in that study. The fourth column presents the proportion estimate followed by their confidence intervals. The sixth and seventh columns present, respectively, the weights for the fixed and random models of the meta-analysis.

Figures 5 and 6 (Supplementary Material) present the meta-analysis of the systematic review of studies on the *mtr*R gene. Figure 5 (Supplementary) presents the result of the meta-analysis of studies on reduced susceptibility to the antibiotic cefixime with the proportion of mutations in the *mtr*R gene. The results of the fixed model show that 96% (CI 88% - 99%) of the analyzed isolates had mutations in the *mtr*R gene. Figure 6 (Supplementary) presents the result of the meta-analysis of cefixime-resistant isolates with the proportion of mutations in the *mtr*R gene. The results of the fixed model indicate that 94% (CI 65% - 99%) of the analyzed isolates had mutations in the *mtr*R gene.

**Figure. 5.**
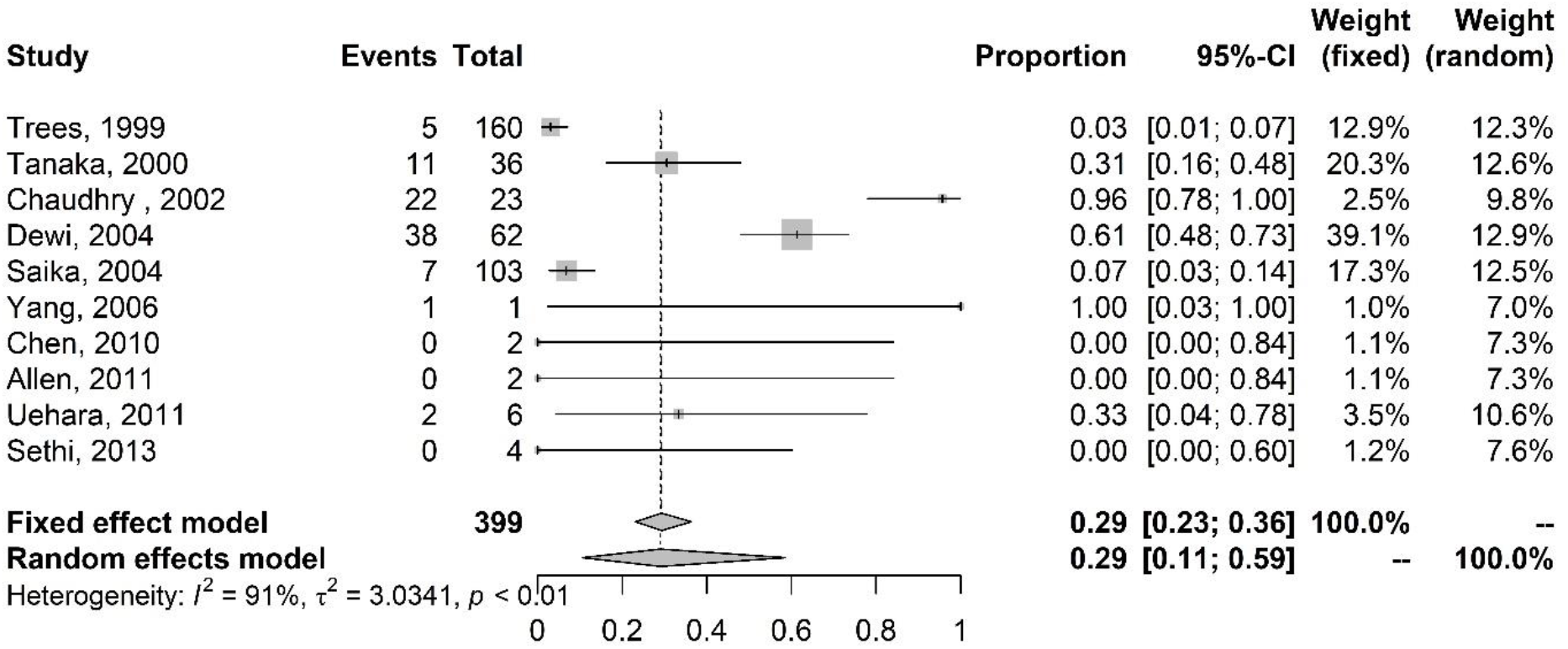
Meta-analysis of the proportion of gene mutations in isolates with intermediate resistance to ciprofloxacin (parC gene). The results are presented in forest plots. In its first column (“study”) are the citations of the selected studies. The second column (“events”) contains the number of isolates with mutations in the evaluated gene, while the third column (“total”) shows the total number of samples analyzed in that study. The fourth column presents the proportion estimate followed by their confidence intervals. The sixth and seventh columns present, respectively, the weights for the fixed and random models of the meta-analysis.

**Figure. 6.**
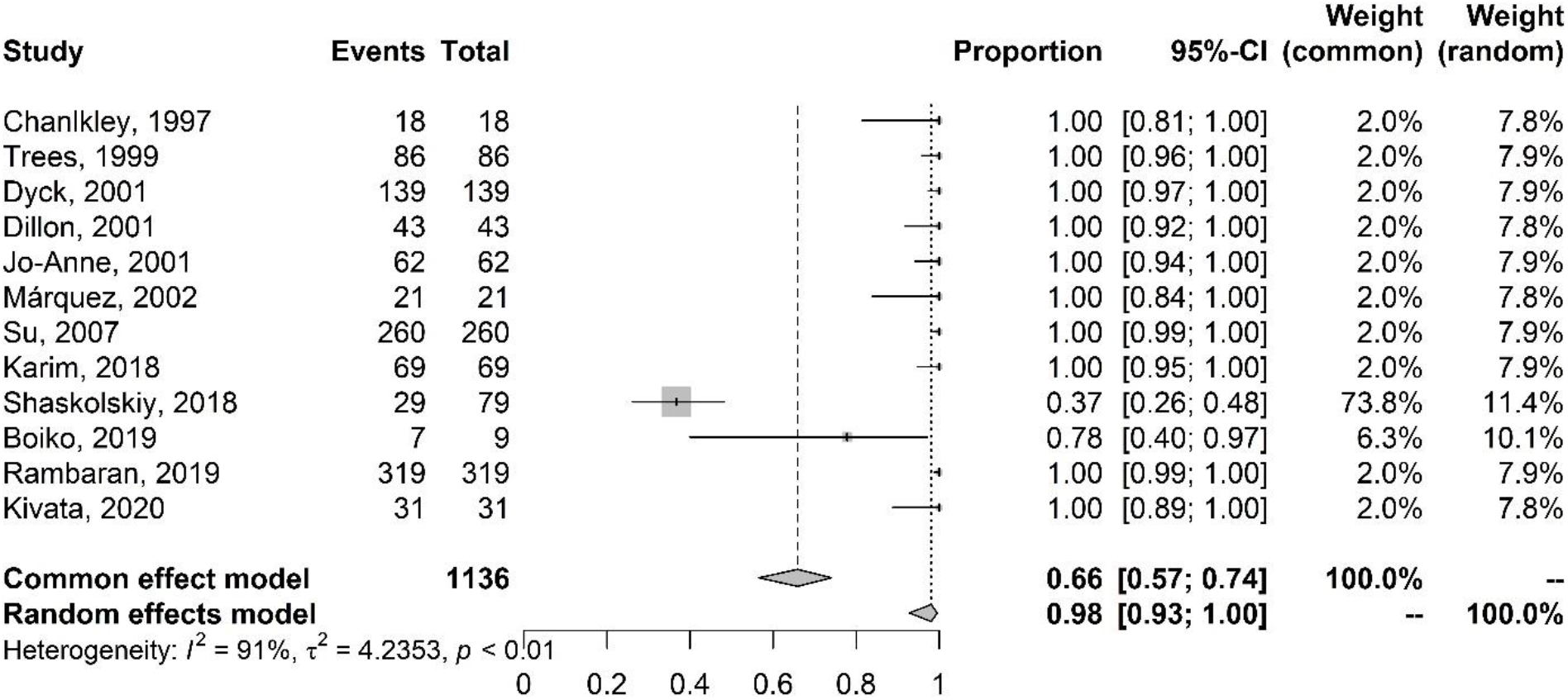
Meta-analysis of the proportion of gene mutations in tetracycline-resistant isolates and the presence of the *tet*M gene. The results are presented in forest plots. In its first column (“study”) are the citations of the selected studies. The second column (“events”) contains the number of isolates with mutations in the evaluated gene, while the third column (“total”) shows the total number of samples analyzed in that study. The fourth column presents the proportion estimate followed by their confidence intervals. The sixth and seventh columns present, respectively, the weights for the fixed and random models of the meta-analysis.

### 3.4 Meta-analysis of studies evaluating resistance and reduced susceptibility to ceftriaxone

Figure 7 (Supplementary Material) presents the meta-analysis on ceftriaxone resistance with the proportion of mutations in the *pen*A (Figure 7A) and *mtr*R (Figure 7B) genes. The results of the fixed model show that 93% (CI 82% - 97%) of the analyzed isolates had mutations in the *pen*A gene (Figure 7A), and 90% (CI 72% - 97%) of the isolates had mutations in *mtr*R (Figure 7B).

Figure 8 (Supplementary Material) shows the meta-analysis of studies on reduced susceptibility to ceftriaxone with the proportion of mutations in the *pen*A (Figure 8A) and *mtr*R (Figure 8B) genes. The results of the random models indicate that 95% (CI 85% -99%) of the isolates had mutations in the *pen*A gene (Figure 8A) and 94% (CI 80% - 99%) of the isolates had mutations in the *mtr*R gene (Figure 8B).

Figures 9 and 10 (Supplementary Material) present the meta-analysis of the systematic review of the *mtr*R gene. Figure 9 (Supplementary) presents the meta-analysis of studies on reduced susceptibility to ceftriaxone with the proportion of mutations in the *mtr*R gene. Results from the random model indicate that 71% (CI 58% - 82%) of the isolates had mutations in the *mtr*R gene.

Figure 10 (Supplementary Material) presents the meta-analysis of studies on ceftriaxone resistance with the proportion of *mtr*R mutations. Fixed effect results show that 93% (CI 72% - 99%) of the isolates had mutations in the *mtr*R gene.

### 3.5 Meta-analysis of studies evaluating resistance to azithromycin

The meta-analysis of studies of azithromycin-resistant isolates with the proportion of mutations in the *mtr*R gene is shown in Figure 1. The results of the fixed model show that 95% (CI 88% - 98%) of the analyzed isolates had mutations in the *mtr*R gene.

### 3.6 Meta-analysis of studies evaluating resistance and intermediate resistance to ciprofloxacin

The meta-analysis of studies on ciprofloxacin resistance with the proportion of mutations in the *gyr*A gene is shown in Figure 2. The results of the randomized model show that 97% (95% -99% CI) of the isolates had mutations in the *gyr*A gene.

Figure 3 presents the meta-analysis of studies on ciprofloxacin resistance with the proportion of mutations in the *par*C gene. The results of the random effect indicate that 88% (CI 83% - 92%) of the analyzed isolates have mutations in the *par*C gene.

Figure 4 presents the results of the meta-analysis of studies on intermediate resistance to ciprofloxacin with the proportion of mutations in the *gyr*A gene. The results of the random model indicate that 91% (CI 78% - 97%) of the isolates had mutations in the *gyr*A gene.

The meta-analysis of studies on intermediate resistance to ciprofloxacin with the proportion of mutations in the *par*C gene is shown in Figure 5. Random effect results indicate that 29% (CI 11% - 59%) of the isolates had mutations in the *par*C gene.

### 3.7 Meta-analysis of studies evaluating tetracycline resistance

The presence of the plasmid containing the *tet*M gene and tetracycline resistance was also evaluated. Figure 6 presents the result of the meta-analysis of studies on the antibiotic tetracycline with the proportion of plasmids containing the *tet*M gene. The random effect results indicate that 98% of the analyzed isolates had the *tet*M gene.

## 4. DISCUSSION

The present study estimated the frequency of mutations in the genes of *N. gonorrhoeae* isolates conferring resistance to the antibiotics penicillin, cefixime, ceftriaxone, tetracycline, azithromycin and ciprofloxacin. As shown by the results obtained in this study, researchers in several countries have studied mechanisms of resistance of *N. gonorrhoeae* to antibiotics. It is evident that *N. gonorrhoeae* has developed or acquired resistance to several classes of antibiotics. Here, it was possible to determine the existence of resistance and/or reduced susceptibility to penicillin, ceftriaxone, cefixime, tetracycline, azithromycin and ciprofloxacin and show that mutations in genes encoding target proteins of these antibiotics are present in more than 50% of the isolates. Cases with elevated MICs for third-generation cephalosporins have been reported worldwide. These isolates are of concern, as these antibiotics are the last option for treating gonorrhoea.

The results of meta-analyses of the *pen*A and *mtr*R genes revealed that there is a relationship between reduced susceptibility and/or resistance to penicillin, ceftriaxone and cefixime and mutations in these genes in different countries over time. In addition, the meta-analyses of studies on the *gyr*A and *par*C genes showed that mutations in these genes are involved in ciprofloxacin resistance. Moreover, the presence of a plasmid containing the *tet*M gene is related to tetracycline resistance, as shown by the meta-analysis of studies on this gene. The results also showed that mutations in the *mtr*R gene are involved in the development of resistance to cefixime, ceftriaxone and azithromycin. The relationship between the presence of mutations and antibiotic resistance was supported by the presence of mutations in more than 50% of isolates with resistance, intermediate resistance or reduced susceptibility.

The studies also evaluated the main mutations found (Supplementary). The most common mutations in *pen*A contribute to resistance to β-lactams. The A501V, A501T and G545S mutations may increase the MICs across the spectrum of cephalosporins (14). Mosaic-like structures in PBP2 were found in several isolates (Tables 1 and 2, column 3). Meanwhile, Takahata et al. (2006) (15) found that the presence of mosaicism in PBP2 was strongly associated with reduced susceptibility to cefixime and other cephalosporins. Commensal *Neisseria* are reservoirs of resistance genes and are linked to the increase in *N. gonorrhoeae* resistant isolates. Takahata et al. (2006) (15) proposed that horizontal transfer of *pen*A genes among *Neisseria* resulted in allelic mosaicism in *N. gonorrhoeae* and *N. meningitidis*. However, some alterations such as G545S are the result of selective pressure by antibiotics.

Mutations in *mtr*R, such as the deletion of alanine (A) in the 13 bp inverted repeat sequence in the promoter region and more common substitutions such as G45D in the coding region, were shown to cause the overexpression of MtrCDE efflux pumps and the increase in efflux. These were in turn revealed to be related to resistance to penicillin and azithromycin and reduced susceptibility to cefixime and ceftriaxone (14). The efflux pump system is one of the essential factors behind multi-drug resistance (16). The presence of other mutations such as in *mtr*R may be necessary for the high MIC level of ceftriaxone (17).

Meta-analysis results of the *gyr*A and *par*C genes showed that there is a higher proportion of mutations in *gyr*A than in *par*C, mainly in isolates with intermediate resistance. Costa-Lourenço et al. (2017) (18) suggested that mutations in *gyr*A are more important for resistance to the quinolone ciprofloxacin as, in most cases, there is no mutation only in *par*C. In addition, Yang et al. (2006) (19) suggested that mutations in *gyr*A determine the resistance in *N. gonorrhoeae*, while mutations in *par*C are related to a high level of resistance. The most common mutations in *gyr*A, such as S91F and D95N, decreased the binding of fluoroquinolones to the DNA gyrase subcomponent, while changes in *par*C such as D86N and S88P decreased the binding of fluoroquinolones to the topoisomerase IV subcomponents (8).

One of the characteristics of the studies that drew attention is the greater number of samples collected from male individuals in the studies that provide information on the sex of the subjects. Given that men are generally more symptomatic than women (3), the demand for care by men is greater, which explains the greater amount of information related to this group. Data on sex, sexual behaviour and age may be relevant given that there is a higher incidence of gonorrhoea cases in particular populations, such as men who have sex with men, sex workers and young adults. Highlighting these data may be important to achieve a more complete analysis of the epidemiology of infection. However, few studies have reported this information. The incompleteness of the data makes it difficult to assess the quality of studies in the literature. The STROBE (Strengthening the reporting of observational studies in epidemiology) statement (20) has recommendations that can improve the quality of epidemiological research reports.

Notably, there is an abundance of studies from Asia, such as China and Japan, but limited research from Latin American and Africa. The lack of Latin American and African epidemiological studies makes it difficult to monitor the infection and the advance of antibiotic resistance in these regions, while also affecting the clinical management of infections there.

The results also demonstrate that the relevant mutations are consistent over time and between different countries and regions. These epidemiological analyses are essential for the control of gonorrhoea, as measures to prevent the spread can be focused on the populations of interest. Studies on mutations and their connection with antibiotic resistance are essential for monitoring the infection.

Several countries reported lower effectiveness of azithromycin and ceftriaxone. Given that *N. gonorrhoeae* is naturally competent for transformation, as with other *Neisseria*, monitoring resistant isolates and adequate clinical management for gonorrhoea are essential. In addition, inadequate clinical management for other bacterial infections also accelerates the increase in resistance, favoring the transfer of resistance genes among bacteria. The emergence and spread of antibiotic-resistant *N. gonorrhoeae* may result in irreversible intractable gonorrhoea.

## CONCLUSION

The development of resistance of gonorrhoea to various antibiotics has led to changes in treatment over the years. Genetic studies, which identify the mechanisms of resistance, and studies on pathology, evolution, and epidemiology are necessary to establish adequate treatment. Moreover, such studies in combination with the monitoring of transmission and increased resistance may even contribute to the development of new antibioticsFurthermore, until an effective treatment or vaccine for gonorrhoea is developed, activities to contain its spread and slow its rise of resistance are essential. These activities include prevention, early diagnosis, epidemiological monitoring, adequate treatment and awareness programs. The results of this study highlight the urgency of continuously monitoring these resistance genes. Putting these measures into practice will positively affect the quality of life of infected individuals and reduce the financial burden on healthcare systems.

## Supporting information

Full Supplementary Data and Tables

## Data Availability

All data produced in the present study are available upon reasonable request to the authors.

## Acknowledgments

The authors wish to thank the following funding sources: FAPEMIG (PPM-00604-16), CNPq (404182/2016-0), and CAPES. DB is a recipient of CNPq fellowship (311876/2020-0). The authors also thank BioMed Proofreading for English language editing of this manuscript.

