## Supplementary material for "The frequency of mutations in the *pen*A, *mtr*R, *gyr*A and *par*C genes of *Neisseria gonorrhoeae*, the presence of *tet*M gene and antibiotic resistance/susceptibility: a systematic review and meta-analyses": Full Supplementary Data and Tables

Running head: *N. gonorrhoeae and antibiotic resistance*

#Address correspondence to Diana Bahia, or Renan P. Souza,. Departamento de Genética, Ecologia e Evolução- GEE, Instituto de Ciências Biológicas, Universidade Federal de Minas Gerais. Av. Antônio Carlos 6627, Pampulha, 31270-901, Caixa Postal 486, Belo Horizonte, MG, Brazil. Tel. 55-31-3409-2568; FAX: 55-31-3409-2567.

### SUPPLEMENTARY MATERIAL

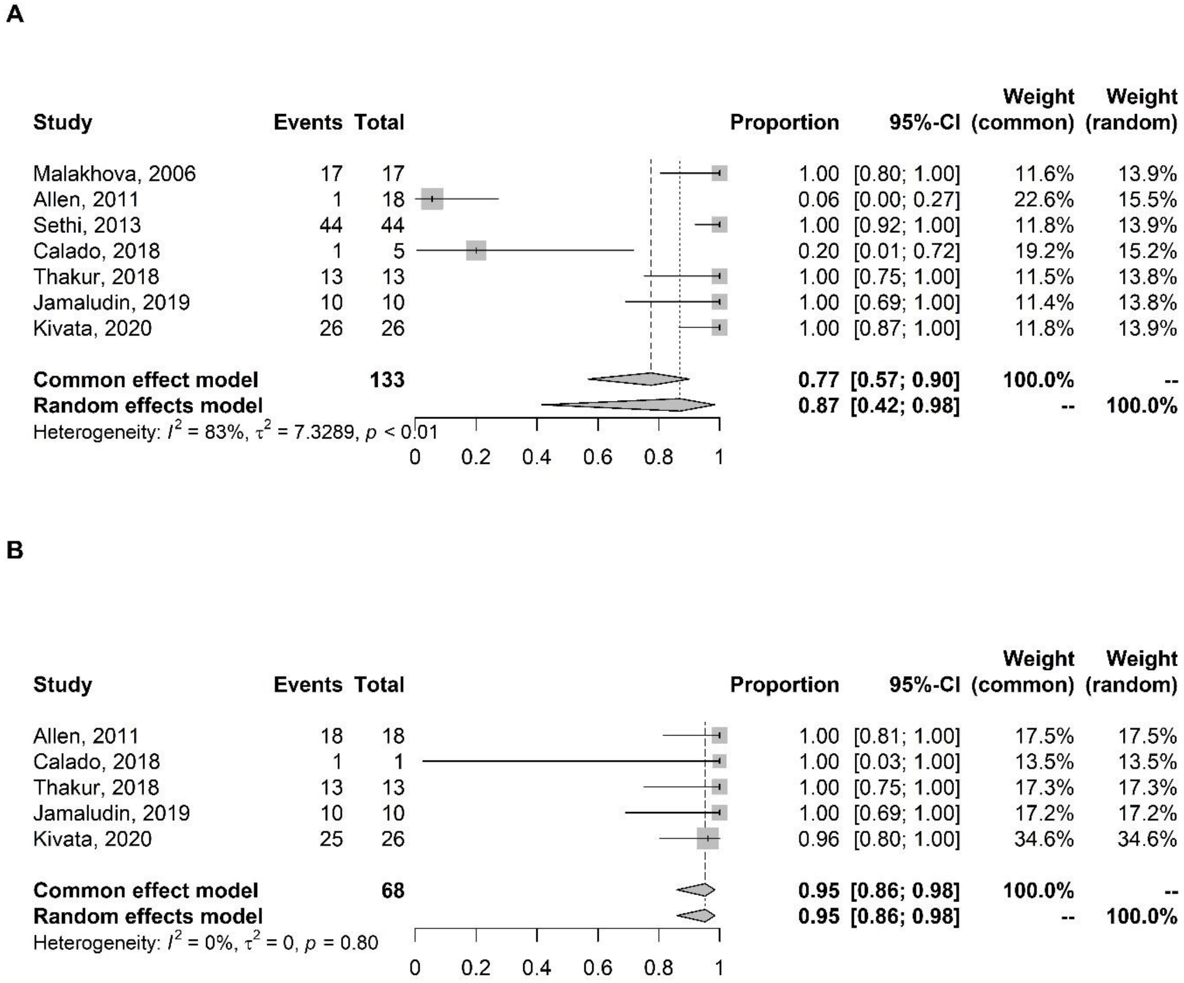


**Figure 1.** Meta-analysis of the proportion of gene mutations in isolates resistant to the antibiotic penicillin. (A) *pen*A. (C) *mtr*R. The results are presented in forest plots. In its first column ("study") are the citations of the selected studies. The second column ("events") contains the number of isolates with mutations in the evaluated gene, while the third column ("total") shows the total number of samples analyzed in that study. The fourth column presents the proportion estimate followed by their confidence intervals. The sixth and seventh columns present, respectively, the weights for the fixed and random models of the meta-analysis.

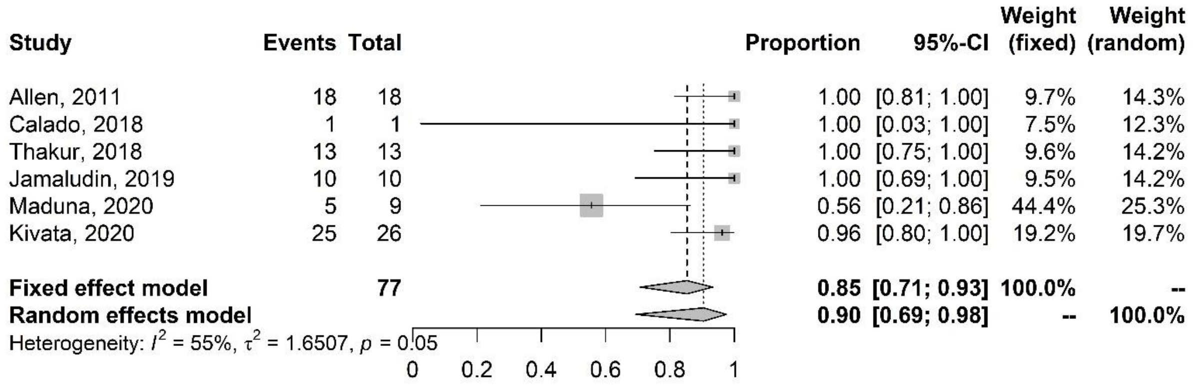


**Figure. 2**. Meta-analysis of the proportion of gene mutations in isolates resistant to penicillin and *mtr*R gene. The results are presented in forest plots. In its first column ("study") are the citations of the selected studies. The second column ("events") contains the number of isolates with mutations in the evaluated gene, while the third column ("total") shows the total number of samples analyzed in that study. The fourth column presents the proportion estimate followed by their confidence intervals. The sixth and seventh columns present, respectively, the weights for the fixed and random models of the meta-analysis.

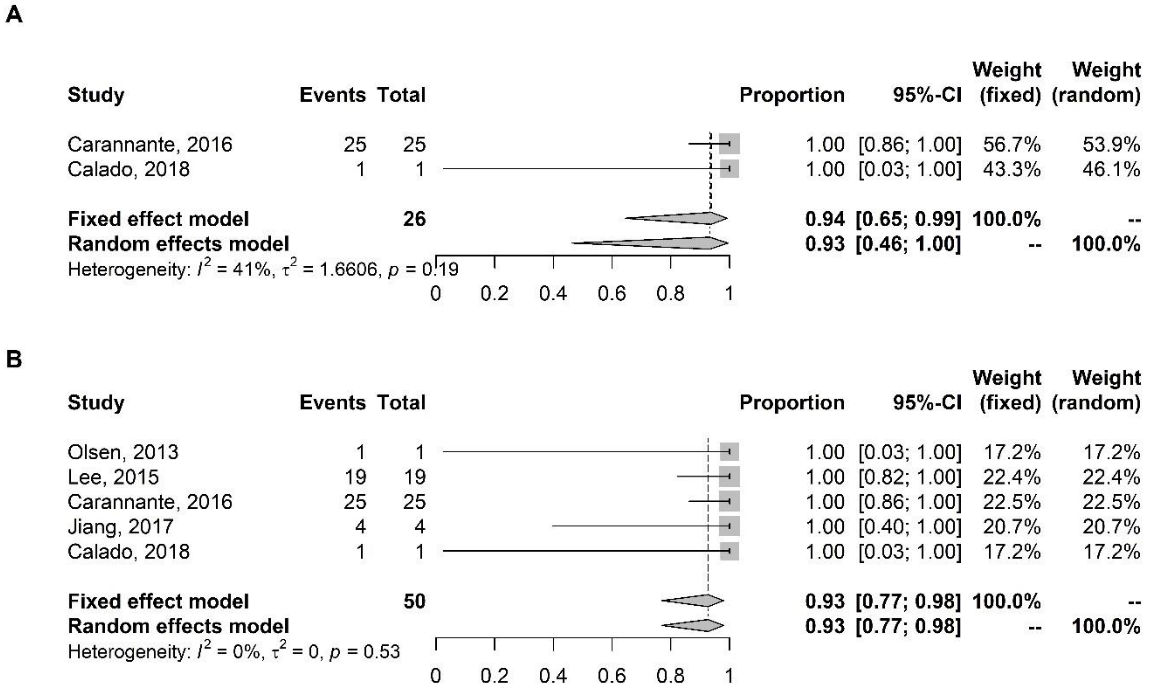


**Figure. 3.** Meta-analysis of the proportion of gene mutations in cefixime-resistant isolates. (A) *mtr*R. (B) *pen*A. The results are presented in forest plots. In its first column ("study") are the citations of the selected studies. The second column ("events") contains the number of isolates with mutations in the evaluated gene, while the third column ("total") shows the total number of samples analyzed in that study. The fourth column presents the proportion estimate followed by their confidence intervals. The sixth and seventh columns present, respectively, the weights for the fixed and random models of the meta-analysis.

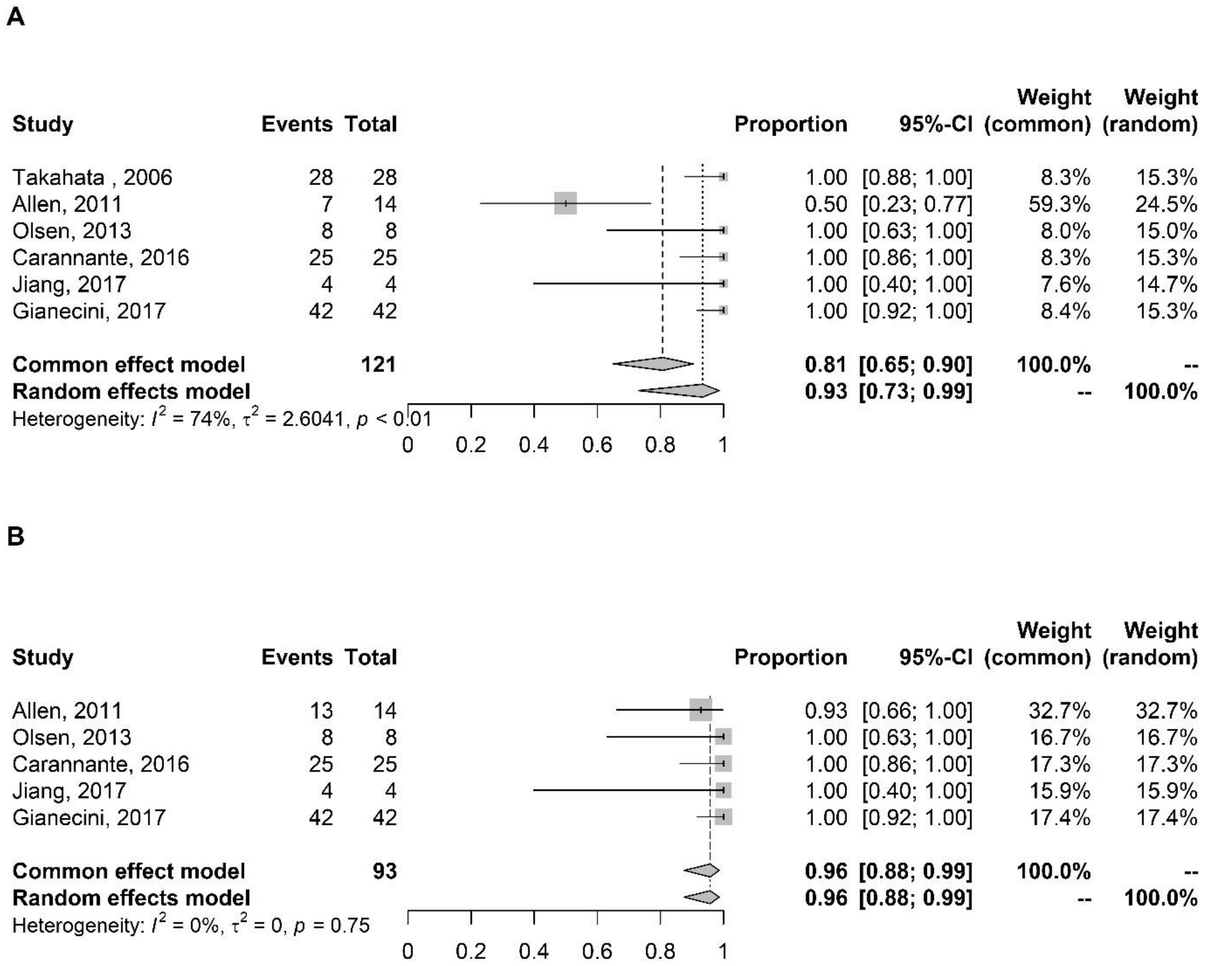


**Figure 4.** Meta-analysis of the proportion of gene mutations in isolates with reduced susceptibility to cefixime. (A) *pen*A. (B) *mtr*R. The results are presented in forest plots. In its first column ("study") are the citations of the selected studies. The second column ("events") contains the number of isolates with mutations in the evaluated gene, while the third column ("total") shows the total number of samples analyzed in that study. The fourth column presents the proportion estimate followed by their confidence intervals. The sixth and seventh columns present, respectively, the weights for the fixed and random models of the meta-analysis.

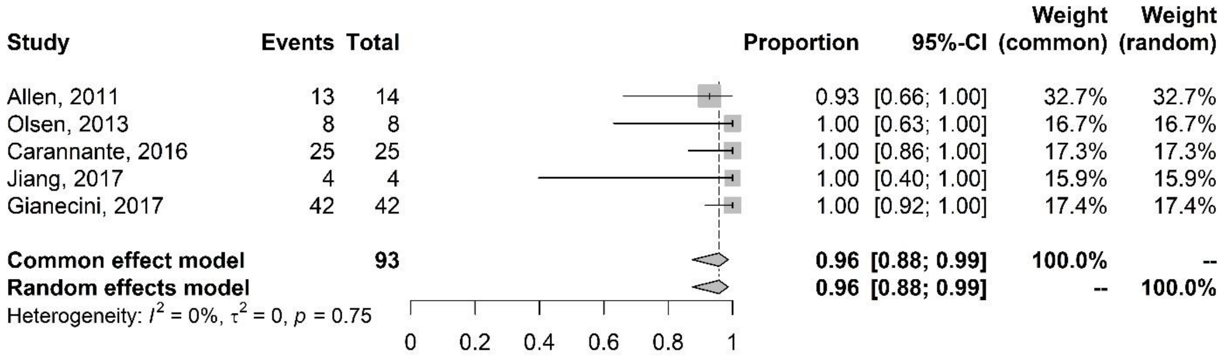


**Figure. 5.** Meta-analysis of the proportion of gene mutations in isolates with reduced susceptibility to cefixime (*mtr*R gene). The results are presented in forest plots. In its first column ("study") are the citations of the selected studies. The second column ("events") contains the number of isolates with mutations in the evaluated gene, while the third column ("total") shows the total number of samples analyzed in that study. The fourth column presents the proportion estimate followed by their confidence intervals. The sixth and seventh columns present, respectively, the weights for the fixed and random models of the meta-analysis.

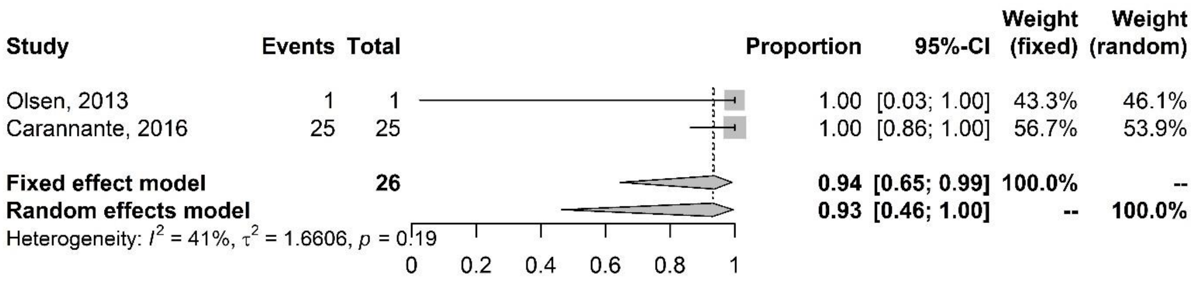


**Figure. 6.** Meta-analysis of the proportion of gene mutations in cefixime resistant isolates (*mtr*R gene). The results are presented in forest plots. In its first column ("study") are the citations of the selected studies. The second column ("events") contains the number of isolates with mutations in the evaluated gene, while the third column ("total") shows the total number of samples analyzed in that study. The fourth column presents the proportion estimate followed by their confidence intervals. The sixth and seventh columns present, respectively, the weights for the fixed and random models of the meta-analysis.

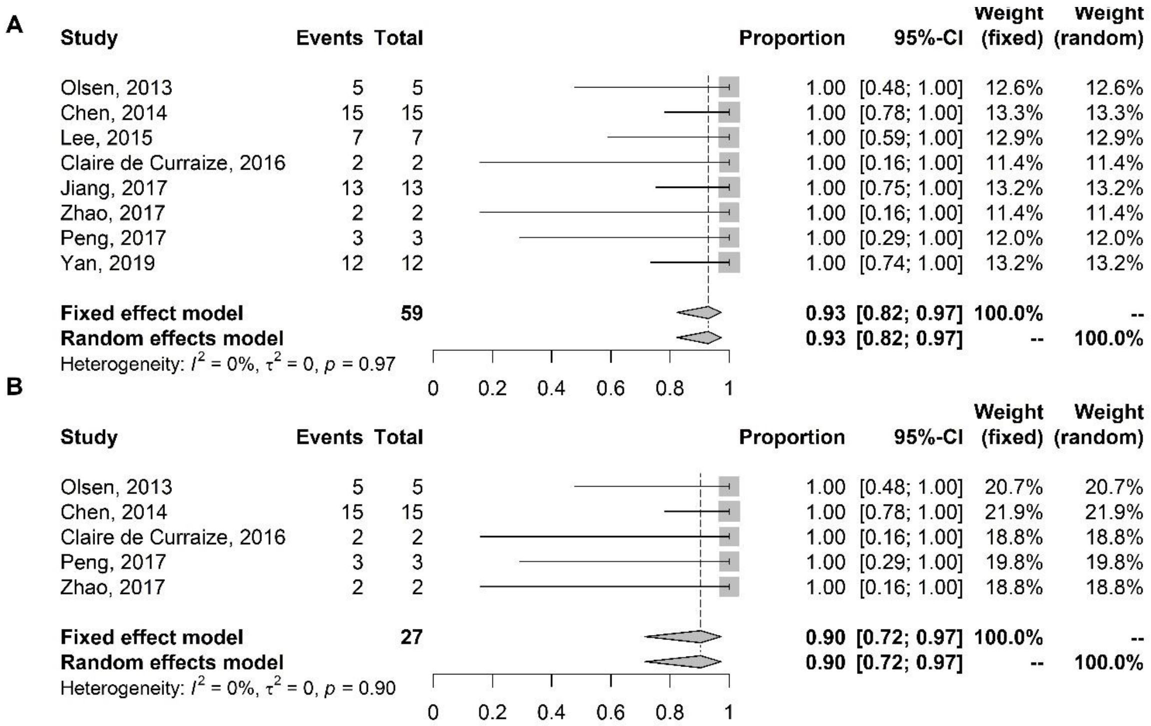


**Figure. 7.** Meta-analysis of the proportion of gene mutations in ceftriaxone-resistant isolates. (A) *pen*A. (B) *mtr*R. The results are presented in forest plots. In its first column ("study") are the citations of the selected studies. The second column ("events") contains the number of isolates with mutations in the evaluated gene, while the third column ("total") shows the total number of samples analyzed in that study. The fourth column presents the proportion estimate followed by their confidence intervals. The sixth and seventh columns present, respectively, the weights for the fixed and random models of the meta-analysis.

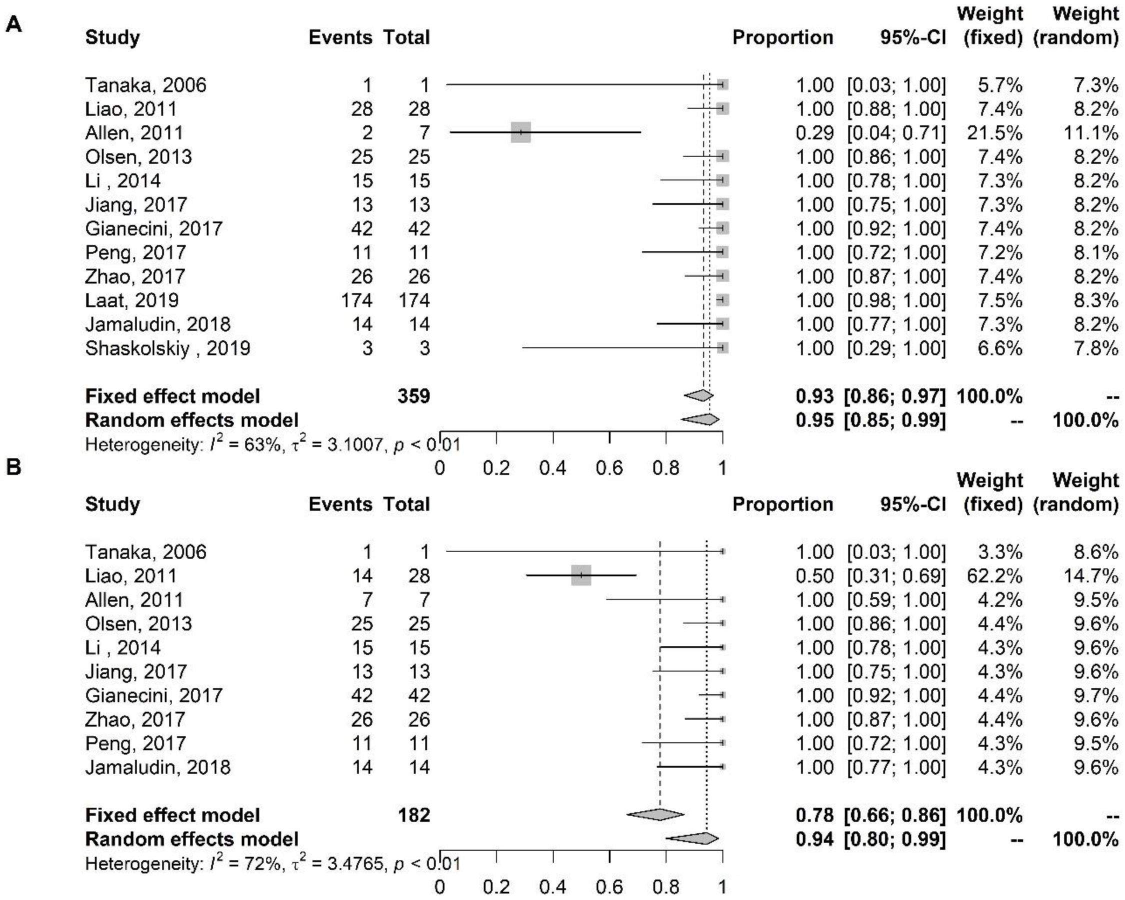


**Figure. 8.** Meta-analysis of the proportion of gene mutations in isolates with reduced susceptibility to ceftriaxone. (A) *pen*A. (B) *mtr*R. The results are presented in forest plots. In its first column ("study") are the citations of the selected studies. The second column ("events") contains the number of isolates with mutations in the evaluated gene, while the third column ("total") shows the total number of samples analyzed in that study. The fourth column presents the proportion estimate followed by their confidence intervals. The sixth and seventh columns present, respectively, the weights for the fixed and random models of the meta-analysis.

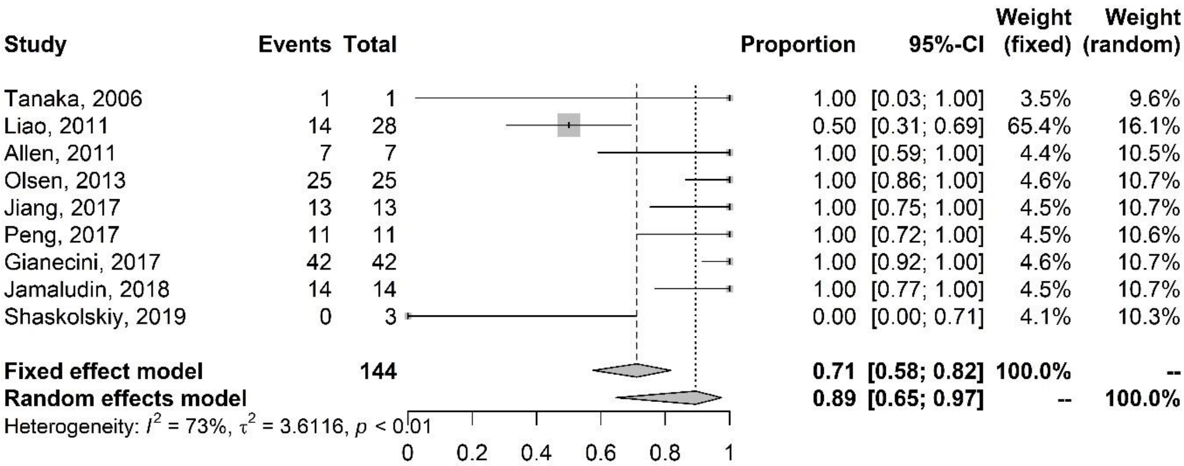


**Figure. 9.** Meta-analysis of the proportion of gene mutations in isolates with reduced susceptibility to ceftriaxone (*mtr*R gene). The results are presented in forest plots. In its first column ("study") are the citations of the selected studies. The second column ("events") contains the number of isolates with mutations in the evaluated gene, while the third column ("total") shows the total number of samples analyzed in that study. The fourth column presents the proportion estimate followed by their confidence intervals. The sixth and seventh columns present, respectively, the weights for the fixed and random models of the meta-analysis.

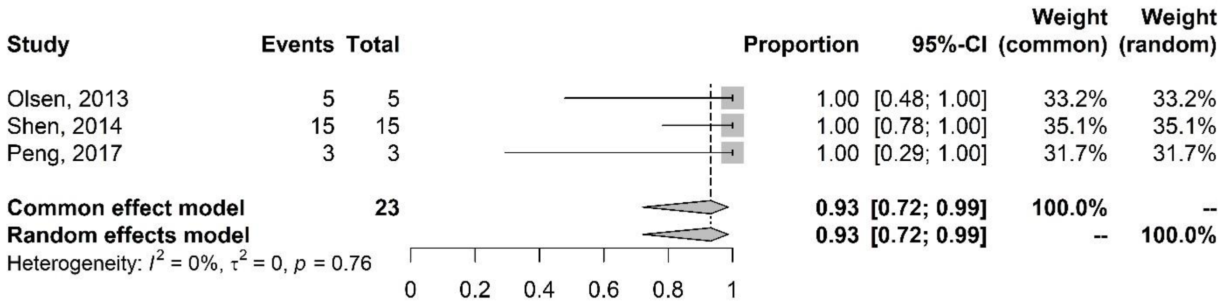


**Figure. 10.** Meta-analysis of the proportion of gene mutations in isolates resistant to ceftriaxone (*mtr*R gene). The results are presented in forest plots. In its first column ("study") are the citations of the selected studies. The second column ("events") contains the number of isolates with mutations in the evaluated gene, while the third column ("total") shows the total number of samples analyzed in that study. The fourth column presents the proportion estimate followed by their confidence intervals. The sixth and seventh columns present, respectively, the weights for the fixed and random models of the meta-analysis.

| Author | Year of publication | Study period | Local | Number of isolates | Number of isolates screened | Antibiotics | Gene | M | W | MSM | MSW | SW | Men age | Women age | Anatomic site |
|---|---|---|---|---|---|---|---|---|---|---|---|---|---|---|---|
| Takahata | 2006 | 2002 | Japan | 58 | 58 | Cefixme | *pon*A, *pen*A, *por* | NA | NA | NA | NA | NA | NA | NA | Urethra |
| Tanaka | 2006 | 2000 - 2001 | Japan | 398 | 398 | Ceftriaxone | *pen*A, *pon*A, *mtr*R | NA | NA | NA | NA | NA | NA | NA | Urethra |
| Liao | 2011 | 2005 - 2008 | China | 230 | 230 | Ceftriaxone | *pen*A, *mtr*R, *por* | 230 | 0 | NA | NA | NA | NA | NA | NA |
| Allen | 2011 | 2008 | Canada | 149 | 149 | Ceftriaxone, Cefixime, Penicillin | *pen*A, *pon*A, *mtr*R, *por* | NA | NA | NA | NA | NA | NA | NA | NA |
| Olsen | 2013 | 2011 | Vietnam | 108 | 108 | Ceftriaxone, Cefixime | *pen*A, *mtr*R, *por*B | 85 | 19 | NA | NA | NA | 28.5 | 29 | NA |
| Li | 2014 | 2011 e 2012 | China | 340 | 334 | Cefixime | *pen*A, *mtr*R, *por*B, *pon*A | 340 | NA | NA | NA | NA | NA | NA | Urethra |
| Carannante | 2016 | 2011 - 2014 | Italy | 900 | 65 | Cefixime | *pen*A, *mtr*R | NA | NA | NA | NA | NA | NA | NA | NA |
| Jiang | 2017 | 2014 - 2015 | China | 126 | 126 | Cefixime, Ceftriaxone | *pen*A, *mtr*R | NA | NA | NA | NA | NA | NA | NA | Uretrha, cervice |
| Peng | 2017 | 2015 - 2016 | China | 128 | 128 | Ceftriaxone | *pen*A, *mtr*R, *pen*B | 120 | 8 | NA | NA | NA | 32,8±10,1 | 34,8 +- 9,4 | Urethra, endocervice |
| Gianecini | 2017 | 2009 - 2013 | Argentina | 1987 | 1987 | Ceftriaxone, Cefixime | *pen*A, *mtr*R | NA | NA | NA | NA | NA | NA | NA | NA |
| Jamaludin | 2018 | 2012 - 2015 | New Zealand | 28 | 28 | Penicillin | *pen*A, *mtr*R, *pen*B, *pon*A | NA | NA | NA | NA | NA | NA | NA | NA |
| De Laat | 2019 | 2009 - 2017 | Netherlands | 348 | 348 | Ceftriaxone | *pen*A | 320 | 28 | 314 | 6 | 15 | NA | NA | Urethra, rectum, cervice, pharynx |
| Shaskolskiy | 2019 | 2015 - 2017 | Russia | 522 | 522 | Penicillin, Ceftriaxone | *pen*A | NA | NA | NA | NA | NA | NA | NA | NA |

**Table 1.** Characteristic of studies of isolates with reduced susceptibility to ceftriaxone and cefixime. M= men. W= women. MSM = men who have sex with men. MSW = men who have sex with women. SW= sex workers. NA = not available.

| Author | Year of publication | Study period | Local | Number of isolates | Number of isolates screened | Antibiotics | Gene | M | W | MSM | MSW | SW | Men age | Women age | Anatomic site |
|---|---|---|---|---|---|---|---|---|---|---|---|---|---|---|---|
| Malakhova | 2006 | NA | Russia | 31 | 31 | Penicillin | *pen*A, *pon*A, *pen*B | NA | NA | NA | NA | NA | NA | NA | NA |
| Allen | 2011 | 2008 | Canada | 149 | 149 | Ceftriaxone, Cefixime, Penicillin | *pen*A, *pon*A, *mtr*R, *por* | NA | NA | NA | NA | NA | NA | NA | NA |
| Olsen | 2013 | 2011 | Vietnam | 108 | 108 | Ceftriaxone, Cefixime | *pen*A, *mtr*R, *por* | 85 | 19 | NA | NA | NA | 28.5 | 29 | NA |
| Chen | 2014 | 2007- 2012 | China | 278 | 15 | Ceftriaxone | *pen*A, *mtr*R, *pen*B | 232 | 46 | NA | NA | NA | NA | NA | Urethra, endocervice |
| Lee | 2015 | 2011 - 2013 | Korea | 210 | 210 | Ceftriaxone, Cefixime | *pen*A | 136 | 47 | NA | NA | 27 | NA | NA | NA |
| Claire de Curraize | 2016 | 2010 - 2014 | France | 6340 | 6340 | Ceftriaxone | *pen*A, *mtr*R, *pen*B | NA | NA | NA | NA | NA | NA | NA | NA |
| Carannante | 2016 | 2011 - 2014 | Italy | 900 | 65 | Cefixime | *pen*A, *mtr*R | NA | NA | NA | NA | NA | NA | NA | NA |
| Peng | 2017 | 2015 - 2016 | China | 128 | 128 | Ceftriaxone | *pen*A, *mtr*R, *pen*B | 120 | 8 | NA | NA | NA | 32,8 ± 10,1 | 34,8 ± 9,4 | Urethra, endocervice |
| Thakur | 2018 | 2003 - 2008 | Canada | 146 | 146 | Penicillin | *pen*A, *mtr*R, *pen*B | NA | NA | NA | NA | NA | NA | NA | NA |
| Jamaludin | 2018 | 2012 - 2015 | New Zaeland | 28 | 28 | Penicillin | *pen*A, *mtr*R, *pen*B, *pon*A | NA | NA | NA | NA | NA | NA | NA | NA |
| Yan | 2019 | 2015 - 2017 | China | 379 | 379 | Ceftriaxone | *pen*A | NA | NA | NA | NA | NA | NA | NA | NA |
| Calado | 2019 | 2013 - 2015 | Portugal | 30 | 30 | Penicillin, Cefixime | *pen*A, *mtr*R | 30 | 0 | 30 | 0 | NA | NA | NA | Urethra, anus |
| Kivata | 2020 | 2013 - 2018 | Kenya | 36 | 36 | Penicillin | *pen*A, *pen*B, *mtr*R, *pon*A | NA | NA | NA | NA | NA | NA | NA | Urethra, endocevice |

**Table 2.** Characteristics of studies of isolates resistant to penicillin, ceftriaxone, and cefixime. M= men. W= women. MSM = men who have sex with men. MSW = men who have sex with women. SW= sex workers. NA = not available.

| Author | Year of publication | Study period | Local | Number of isolates | Number of isolates analisados | Antibiotics | Gene | M | W | MSM | MSW | SW | Men age | Women age | Anatomic site |
|---|---|---|---|---|---|---|---|---|---|---|---|---|---|---|---|
| Tanaka | 2006 | 2000 a 2001 | Japan | 398 | 398 | Ceftriaxone | *pen*A, *pon*A, *mtr*R | NA | NA | NA | NA | NA | NA | NA | Urethra |
| Liao | 2011 | 2005 e 2008 | China | 230 | 230 | Ceftriaxone | *pen*A, *mtr*R, *por* | 230 | 0 | NA | NA | NA | NA | NA | NA |
| Allen | 2011 | 2008 | Canada | 149 | 149 | Ceftriaxone, Cefixime, Penicillin | *pen*A, *pon*A, *mtr*R, *por* | NA | NA | NA | NA | NA | NA | NA | NA |
| Olsen | 2013 | 2011 | Vietnam | 108 | 108 | Ceftriaxone, Cefixime | *pen*A, *mtr*R, *por*B | 85 | 19 | NA | NA | NA | 28.5 | 29 | NA |
| Jiang | 2017 | 2014 e 2015 | China | 126 | 126 | Cefixime, Ceftriaxone, | *pen*A, *mtr*R | NA | NA | NA | NA | NA | NA | NA | Urethra, cervice |
| Peng | 2017 | 2015 - 2016 | China | 128 | 128 | Ceftriaxone | *pen*A, *mtr*R, *pen*B | 120 | 8 | NA | NA | NA | 32,8 ± 10,1 | 34,8 ± 9,4 | Urethra endocervice |
| Gianecini | 2017 | 2009 - 2013 | Argentina | 1987 | 1987 | Ceftriaxone, Cefixime | *pen*A, *mtr*R | NA | NA | NA | NA | NA | NA | NA | NA |
| Carannante | 2016 | 2011 - 2014 | Italy | 900 | 65 | Cefixime | *pen*A, *mtr*R | NA | NA | NA | NA | NA | NA | NA | NA |
| Jamaludin | 2018 | 2012 - 2015 | New Zaeland | 28 | 28 | Penicillin | *pen*A, *mtr*R, *pen*B, *pon*A | NA | NA | NA | NA | NA | NA | NA | NA |
| Shaskolskiy | 2019 | 2015-2017 | Russia | 522 | 522 | Penicillin, Ceftriaxone | *pen*A | NA | NA | NA | NA | NA | NA | NA | NA |

**Table 3.** Characteristic of studies on the *mtr*R gene of isolates with reduced susceptibility. M= men. W= women. MSM = men who have sex with men. MSW = men who have sex with women. SW= sex workers. NA = not available.

| Author | Year of publication | Study period | Local | Number of isolates | Number of isolates analisados | Antibiotics | Gene | M | W | MSM | MSW | SW | Men age | Women age | Anatomic site |
|---|---|---|---|---|---|---|---|---|---|---|---|---|---|---|---|
| Calado | 2019 | 2013 - 2015 | Portugal | 30 | 30 | Penicillin, Cefixime | *penA, mtrR* | 30 | 0 | 30 | 0 | NA | NA | NA | Urethra, anus |
| Maduna | 2020 | NA | South Africa | 51 | 27 | Penicillin, azithromicin, tetracyclina | mtrR | 51 | 0 | 32 | 19 | NA | 27 | NA | Urethra |
| Shen | 2014 | 2007 - 2012 | China | 278 | 278 | Ceftriaxone | *mtrR, penA* | 232 | 46 | NA | NA | NA | 36.4 | 35,2 | Urethra, cervice |
| Olsen | 2013 | 2011 | Vietnam | 108 | 108 | Ceftriaxone, Cefixime | *penA, mtrR, porB* | 85 | 19 | NA | NA | NA | 28.5 | 29 | NA |
| Belkacem | 2016 | 2013 - 2014 | France | 970 | 970 | Azithromicin | *mtr*R | NA | NA | NA | NA | NA | NA | NA | NA |
| Thakur | 2018 | 2003 - 2008 | Canada | 146 | 146 | Penicillin | *penA, mtrR, penB* | NA | NA | NA | NA | NA | NA | NA | NA |
| Peng | 2017 | 2015 - 2016 | China | 128 | 128 | Ceftriaxone | *penA, mtrR, penB* | 120 | 8 | NA | NA | NA | 32.8 +- 10.1 | 34,8 ± 9,4 | Urethra, endocervice |
| Liu | 2019 | 2001 - 2018 | Taiwan | 598 | 598 | Azithromicin | *mtr*R | NA | NA | NA | NA | NA | NA | NA | Urethra, vagina, pus, eye, blood, surgical wound, gastric juice, synovial fluid, Bartholin abscess |
| Zheng | 2019 | 2014 - 2017 | China | 55 | 55 | Azithromicin | *mtr*R | 52 | 3 | NA | NA | NA | 32 | 32 | Urethra, NA |
| Carannante | 2016 | 2011 - 2014 | Italy | 900 | 65 | Cefixime | *penA, mtrR* | NA | NA | NA | NA | NA | NA | NA | NA |
| Jamaludin | 2018 | 2012 - 2015 | New Zaeland | 28 | 28 | Penicillin | *penA, mtrR, penB, ponA* | NA | NA | NA | NA | NA | NA | NA | NA |
| Kivata | 2020 | 2013 - 2018 | Kenya | 36 | 36 | Penicillin | *penA, penB, mtrR, ponA, bla* | NA | NA | NA | NA | NA | NA | NA | Urethra, endocevice |

**Table 4.** Characteristic of studies on the *mtr*R gene of resistant isolates. M= men. W= women. MSM = men who have sex with men. MSW = men who have sex with women. SW= sex workers. NA = not available.

| Author | Year of publication | Study period | Local | Number of isolates | Number of isolates screened | Antibiotics | Gene | M | W | MSM | MSW | SW | Men age | Women age | Anatomic site |
|---|---|---|---|---|---|---|---|---|---|---|---|---|---|---|---|
| Trees | 1999 | 1994 - 1996 | Far East and US | 234 | 234 | Ciprofloxacin | *gyrA, parC* | NA | NA | NA | NA | 107 | NA | NA | NA |
| De Neeling | 2000 | 1991 - 1998 | Netherlands | 2352 | 2352 | Ciprofloxacin | *gyrA, parC* | NA | NA | NA | NA | NA | NA | NA | NA |
| Tanaka | 2000 | 1993 - 1997 | Japan | 85 | 85 | Ciprofloxacin | *gyrA, parC* | NA | NA | NA | NA | NA | NA | NA | NA |
| Tanaka | 2000 | 1993 - 1998 | Japan | 502 | 502 | Ciprofloxacin | *gyrA, parC* | 502 | 0 | NA | NA | NA | NA | NA | NA |
| Chaudhry | 2002 | 2000 - 2001 | India | 63 | 63 | Ciprofloxacin | *gyrA, parC* | NA | NA | NA | NA | NA | NA | NA | Urethra |
| Alcalá | 2003 | 2000 - 2001 | Spain | 147 | 147 | Ciprofloxacin | *gyrA, parC* | NA | NA | NA | NA | NA | NA | NA | NA |
| Saika | 2004 | 1998 - 1999 | Japan | 332 | 332 | Ciprofloxacin | *gyrA, parC* | 200 | 132 | NA | NA | NA | NA | NA | Urethra vagina |
| Yoo | 2004 | 1999 - 2003 | Korea | 817 | 817 | Ciprofloxacin | *gyrA, parC* | NA | NA | NA | NA | NA | NA | NA | NA |
| Vereshchagin | 2004 | 2002 | Russia | 32 | 32 | Ciprofloxacin | *gyrA, parC* | NA | NA | NA | NA | NA | NA | NA | NA |
| Dewi | 2004 | 1999 - 2001 | Japan | 131 | 131 | Ciprofloxacin | *gyrA, parC* | 131 | 0 | NA | NA | NA | NA | NA | NA |
| Uthman | 2004 | 1999 - 2002 | France | 104 | 104 | Ciprofloxacin | *gyrA, parC* | NA | NA | NA | NA | NA | NA | NA | NA |
| Giles | 2004 | 2000 -2001 | Israel | 80 | 80 | Ciprofloxacin | *gyrA, parC* | 53 | 27 | 53 | NA | 27 | NA | NA | pharynx, |
| Yang | 2006 | 2004 - 2005 | China | 159 | 103 | Ciprofloxacin | *gyrA, parC* | 159 | NA | NA | NA | NA | NA | NA | NA |
| Wang | 2006 | 2003 | China | 95 | 54 | Ciprofloxacin | *gyrA, parC* | 78 | 17 | | NA | NA | 33-96 | 33-96 | NA |
| Ilina | 2008 | 2004 - 2005 | Russia | 464 | 464 | Ciprofloxacin | *gyrA, parC* | NA | NA | NA | NA | NA | NA | NA | NA |
| Chen | 2010 | 1999 - 2004 | Taiwan | 90 | 45 | Ciprofloxacin | *gyrA, parC* | NA | NA | NA | NA | NA | NA | NA | NA |
| Allen | 2011 | 2008 | Canada | 149 | 149 | Ciprofloxacin | *gyrA, parC* | NA | NA | NA | NA | NA | NA | NA | NA |
| Uehara | 2011 | 2005 - 2010 | Brazil | 125 | 125 | Ciprofloxacin | *gyrA, parC* | NA | NA | NA | NA | NA | NA | NA | Urethra, vagina, |
| Kulkarni | 2012 | 2007 - 2009 | India | 64 | 64 | Ciprofloxacin | *gyrA, parC* | 49 | 15 | 2 | NA | 7 | NA | NA | NA |
| Sethi | 2013 | 2007 - 2010 | India,Pakistan,Buthan | 65 | 65 | Ciprofloxacin | *gyrA, parC* | 60 | 5 | NA | NA | NA | NA | NA | NA |
| Calado | 2019 | 2013 - 2015 | Portugal | 30 | 30 | Ciprofloxacin | *gyrA, parC* | 30 | NA | 30 | NA | NA | NA | NA | anus, Urethra |
| Kivata | 2019 | NA | Kenya | 84 | 22 | Ciprofloxacin | *gyrA, parC* | 73 | 11 | NA | NA | NA | NA | NA | NA |
| Boiko | 2019 | 2013 - 2018 | Ukraine | 150 | 150 | Ciprofloxacin | *gyrA, parC* | 130 | 20 | 2 | NA | NA | 29,2 | 28 | NA |

**Table 5.** Characteristic of *gyr*A and *par*C studies. M= men. W= women. MSM = men who have sex with men. MSW = men who have sex with women. SW= sex workers. NA = not available.

| Author | Year of publication | Study period | Local | Number of isolates | Number of isolates screened | Antibiotics | Gene | M | W | MSM | MSW | SW | Men age | Women age | Anatomic site |
|---|---|---|---|---|---|---|---|---|---|---|---|---|---|---|---|
| Chalkley | 1997 | 1994 e 1995 | South Africa | 210 | 210 | Tetraciclina | *tetM* | NA | NA | NA | NA | NA | NA | NA | NA |
| Trees | 1999 | 1994 | Ohio | 591 | 591 | Tetraciclina | *tetM* | NA | NA | NA | NA | NA | NA | NA | NA |
| Dyck | 2001 | 1998 a 1999 | Benin | 170 | 143 | Penicillin, tetraciclina | *tetM* | 0 | 143 | 0 | 0 | 143 | NA | NA | vagina |
| Dillon | 2001 | 1994 - 1996 | Guyana, Saint Vincent, Trinidad | 1686 | 1686 | Penicillin, tetraciclina | *tetM* | NA | NA | NA | NA | NA | NA | NA | NA |
| Jo-Anne | 2001 | 1999 | Manaus/Brazil | 168 | 81 | Tetraciclina | *tetM* | 139 | 29 | NA | NA | NA | NA | NA | Urethra, cérvice |
| Márquez | 2002 | 1996 - 1999 | Uruguay | 181 | 181 | Tetraciclina | *tetM* | NA | NA | NA | NA | NA | NA | NA | NA |
| Su | 2007 | 1999 - 2006 | China | 1208 | 1208 | Tetraciclina | *tetM* | NA | NA | NA | NA | NA | NA | NA | NA |
| Karim | 2018 | 2013 - 2015 | Morocco | 149 | 149 | Tetraciclina | *tetM* | 0 | 149 | 0 | 0 | NA | - | 43,16 | NA |
| Shaskolskiy | 2018 | 2015 - 2017 | Russia | 401 | 401 | Tetraciclina | *tetM* | 322 | 79 | NA | NA | NA | 12 a 60 | 12 a 60 | NA |
| Boiko | 2019 | 2013 - 2018 | Ukraine | 150 | 150 | Tetraciclina | *tetM* | NA | NA | NA | NA | NA | NA | NA | NA |
| Rambaran | 2019 | 2013 - 2014 | South Africa | 319 | 319 | Tetraciclina | *tetM* | 248 | 71 | NA | NA | NA | NA | NA | Urethra, cérvice |
| Kivata | 2020 | 2013 - 2018 | Kenya | 36 | 36 | Tetraciclina | *tetM* | NA | NA | NA | NA | NA | NA | NA | Urethra, cérvice |

**Tabela 6.** Characteristics of studies on the *tet*M gene. M= men. W= women. MSM = men who have sex with men. MSW = men who have sexwith women. SW= sex workers. NA = not available.

| Author | Year of publication | Mutations *pen*A | Mutations *mtr*R |
|---|---|---|---|
| Malakhova | 2006 | D345a | - |
| Allen | 2011 | XXXIV; XXXVII; XXXVIII; XXXV | A deletion; G45D; A39T; R44H |
| Olsen | 2013 | A501T; XVIII | A deletion; H105Y; A39T; G45D; D79N, T86A |
| Chen | 2014 | new1; new6; new8; V; XVII; XIII; A501T; A501V; G42S | A deletion; H105Y; A39T; G45D |
| Lee | 2015 | XIII | A deletion; A39T; |
| Claire de Curraize | 2016 | XXXIV; XXXVI | A deletion |
| Carannante | 2016 | XXXIV; A50IV; IV; XIX; XXXVI; P551L; G542S | A deletion; H105Y; G45D; D19N; T86A; A39T |
| Peng | 2017 | II; XVII; XII; A375T; A501V; G452S; P551S | A deletion; G45D; A39T; A40D |
| Thakur | 2018 | IX; II; XII; | A deletion; G45D; H105Y; A39T |
| Yan | 2019 | X; VII; XII; XIII; XXVII | A deletion; G45D |
| Jamaludin | 2019 | I312M; V316T; N512Y; G545S; A501V; P551S | A deletion; H505Y; G34D; A39T; T86A |
| Calado | 2019 | XXXX | A deletion; |
| Kivata | 2020 | XIV; II; IX; XIX; XXII; | A deletion; G45D; A39T; H205Y; D39N; T86A |

**Table 7.** Mutations in *pen*A, and *mtr*R genes from studies of resistant isolates.

| Author | Year of publication | Mutations penA | Mutations mtrR |
|---|---|---|---|
| Takahata | 2006 | F504L; A510V; N512Y; A516G; A509T; P522V; K555Q; I556V; mosaic-1; mosaic-2 | - |
| Tanaka | 2006 | Mosaico | A deletion; G45A; Y105H |
| Liao | 2011 | A501V; A516G; F404L; P551S; I566V; I556V; K555Q | A deletion; 139T; H105Y; G45D |
| Allen | 2011 | XXXIV; XXXVII; XXXVIII; XXXV | A deletion; G45D; A39T; R44H |
| Olsen | 2013 | A501T; XVIII | A deletion; H105Y; A39T; G45D; D79N, T86A |
| Li | 2014 | XVIII; XLI; XIII; | A deletion |
| Jiang | 2017 | XVII; XII; XXV; V; XXI; XVII; A501V; G4542S | A deletion; G45D; A39T; A40D |
| Gianecini | 2017 | XXXIV; X; V; XII; XIII | A deletion; H105Y |
| Peng | 2017 | II; XVII; XII; A375T; A501V; G452S; P551S | A deletion; G45D; A39T; A40D |
| Zhao | 2017 | XXXIV; XII; XIII; XVIII; XXI; A501V; A502V; D345a | A deletion; A39T; G45D; R44G; L47R; H105F |
| Jamaludin | 2018 | I312M; V316T; N512Y; G545S; A501V; P551S | A deletion; H505Y; G34D; A39T; T86A |
| De Laat | 2019 | XXXIV; A501T | - |
| Shaskolskiy | 2019 | D345a | A deletion; T insertion |

**Table 8.** Mutations in *pen*A, and *mtr*R genes from studies of isolates with reduced susceptibility.

| Author | Year of publication | *mtr*R mutations |
|---|---|---|
| Liao | 2011 | A39T; H105Y; G45D; T86A; |
| Tanaka | 2000 | A deletion; D95D; Y105H |
| Allen | 2011 | A deletion; G45D; A39T |
| Olsen | 2013 | A deletion; A39T; H105Y; D79N; T86A; G45D |
| Chen | 2014 | A deletion; H105Y; 545D; A39T |
| Carannante | 2016 | A deletion; H105Y; G24D; |
| Belkacem | 2016 | A deletion; A39T; R44H; A77P; H105Y; D76A; D79N; F178L; A142S |
| Jiang | 2017 | A deletion; G45D; A39T; A40D |
| Peng | 2017 | A deletion; G45D |
| Gianecini | 2017 | A deletion; H105Y |
| Thakur | 2018 | A deletion; G45D; H105Y; A39T |
| Jamaludin | 2018 | A deletion; G45D A39T; H105Y |
| Zheng | 2019 | A deletion; G45D; |
| Calado | 2019 | A deletion; T insertion; G45D; A39T |
| Liu | 2019 | A deletion; A39T; G45D |
| Shaskolskiy | 2019 | No mutations |
| Maduna | 2020 | A deletion; A39T |
| Kivata | 2020 | A deletion; A39T; G45D; T86A; D79N; H105Y |

**Table 9.** Mutations in the *mtr*R gene.

| Autor | Year of publication | *gyr*A | *par*C |
|---|---|---|---|
| Trees | 1999 | S91F | D86N; E91K; R116L; S87N; G85C |
| De Neeling | 2000 | S91F; D95N; D95G | D86N; S87N; E91K |
| Tanaka | 2000 | S91F; D95N; D95G; | S88P; S87R; E91G |
| Tanaka | 2000 | S91F; A67S; A75S; D95G; D95N; | S88P; R116H; A92G; E91G; E91Q; E91K; |
| Chaudry | 2002 | S91F; D95N; D95G | E91G; F100Y; |
| Alcalá | 2003 | S91F; D95G | D86N; S87R; L131L; Y104Y |
| Saika | 2004 | S91F; D95N; D95G; A92P; D95Y; | D86N; S88P; E91G; S87N; S87I |
| Yoo | 2004 | S91F; D95G; | D86N; S87R; E91G |
| Vereshchagin | 2004 | S91F; D95G; | S87R; E91G |
| Dewi | 2004 | S91F; D95N; D95G | D86N; S87R; S88P |
| Uthman | 2004 | S91F; D95N | D86N; E91G |
| Giles | 2004 | S91F; D95N; D95A; D95G | D86N; S87R; S87N; S88P; E91A; E91G; |
| Yang | 2006 | S91F; A92P; D95A; D95G | S87N; S87R; E91A; E91G |
| Wang | 2006 | S91F; D95A; D95G; D95N | D86N; S87N; E91G; S87I |
| Ilina | 2008 | S91F; D95N; D95A; D95G | D87N |
| Chen | 2010 | S91F; D95N; D95A; D95G | S87R; S87N; E91A; D86N |
| Allen | 2011 | S91F; D95G; D95A | S87R; S87N; E91G; E91Q |
| Uehara | 2011 | S91F; D95G; Q102H | D86N; S87R; E91K; E91Q; A92G |
| Kulkarni | 2012 | S91F; D95N; D95G | E91G |
| Sethi | 2013 | S91F; D95N; D96G | D86N; S87N; S87I; S87R; E91G; E91K; E91Q |
| Calado | 2019 | S91F; D95A; D95G | S87N; S87R; E91G; D86N; |
| Kivata | 2019 | S91F; D95G; D95A | E91G |
| Boiko | 2019 | S91F; D95G | S87R; E91G; S87N; D86N |

**Table 10.** Mutations in the *gyr*A and *par*C gene.
